# Real-time surveillance of international SARS-CoV-2 prevalence using systematic traveller arrival screening

**DOI:** 10.1101/2022.10.12.22280928

**Authors:** Adam J Kucharski, Kiyojiken Chung, Maite Aubry, Iotefa Teiti, Anita Teissier, Vaea Richard, Timothy W Russell, Raphaëlle Bos, Sophie Olivier, Van-Mai Cao-Lormeau

## Abstract

**Background:** Effective COVID-19 response relies on good knowledge of infection dynamics, but owing to under-ascertainment and delays in symptom-based reporting, obtaining reliable infection data has typically required large dedicated local population studies. Although many countries implemented SARS-CoV-2 testing among travellers, interpretation of arrival testing data has typically been challenging because arrival testing data were rarely reported systematically, and pre-departure testing was often in place as well, leading to non-representative infection status among arrivals.

**Methods:** In French Polynesia, testing data were reported systematically with enforced pre-departure testing type and timing, making it possible to adjust for non-representative infection status among arrivals. Combining statistical models of PCR positivity with data on international travel protocols, we reconstructed estimates of prevalence at departure using only testing data from arrivals. We then applied this estimation approach to the USA and France, using data from over 220,000 tests from travellers arriving into French Polynesia between July 2020 and March 2022.

**Findings:** We estimated a peak infection prevalence at departure of 2.8% (2.3-3.6%) in France and 1.1% (0.81-3.1%) in the USA in late 2020/early 2021, with prevalence of 5.4% (4.8-6.1%) and 5.5% (4.6-6.6%) respectively estimated for the Omicron BA.1 waves in early 2022. We found that our infection estimates were a leading indicator of later reported case dynamics, as well as being consistent with subsequent observed changes in seroprevalence over time.

**Interpretation:** As well as elucidating previously unmeasured infection dynamics in these countries, our analysis provides a proof-of-concept for scalable tracking of global infections during future pandemics.

**Funding:** Wellcome (206250/Z/17/Z)

## Introduction

Understanding the true extent of SARS-CoV-2 infection within a population is crucial for effective planning and response. As well as being a leading indicator of subsequent disease, it can help inform the timing and design of control measures both domestically and with respect to internal travel [1]. However, throughout the pandemic, insights into underlying infection dynamics have typically been limited, hindering countries’ ability to respond promptly and proportionately.

In the early stages of 2020, large undetected epidemics in several locations were first identified as a result of infected travellers from these areas being detected in other countries [2,3]. Limited testing availability also meant that infection dynamics had to be estimated from lagged indicators such as hospitalisations, with the number of infections derived from available data on severity [4]. Cross-sectional seroprevalence studies have since provided estimates of the extent of infection within populations, but with a lag and without information on when those infections occurred [5]. Moreover, the roll-out of vaccination and emergence of novel variants means estimation of infection dynamics based on severe outcomes or serological data is becoming more challenging [6]. In some instances, countries have tackled these issues by setting up routine community sampling regardless of symptoms, such as the ONS community infection survey and REACT-1 in the UK [7,8], as well as cohort studies tracking infection in specific workplaces, such as healthcare workers [9]. However, the expense and logistical complexity of such studies has resulted in limited deployment globally.

Despite a lack of studies tracking local infection prevalence, large numbers have been tested regardless of symptoms during the pandemic as a result of traveller screening programmes [10]. Countries have predominately used travel testing as a method of control, with positive individuals having to isolate, as well as being prevented from travelling at all if they test positive before departure. Although some countries with strict restrictions on traveller numbers have reported the number of arriving infections detected over time [11] and data have been reported from brief or small-scale testing programmes [12–14], there has been little systematic long-term data available for global travel testing. Tests have been conducted by different companies and agencies, and in combinations that can include both departure and arrival testing, typically without collation of these tests in consistent databases.

Despite the inconsistent way in which travel testing has generally been reported during the pandemic, such screening presents a unique opportunity for real-time surveillance of infection in multiple countries. Local community infection studies, or analysis of departure testing results, can only provide information on infections within the country conducting the study. In contrast, arrival testing can provide insights into infections among travellers from a range of different locations. However, there are challenges to interpreting arrival test results. If testing is implemented at both departure and arrival, then infections detected among arriving travellers only reflect a subset of all the infected individuals who attempted to travel.

Using data on how test positivity varies over the course of infection, and details of departure and arrival protocols, we developed a model of the travel screening process, and hence used arrival prevalence to reconstruct how many travellers would have tested positive in countries of departure. Incorporating data from French Polynesia, which had systematic arrival screening for SARS-CoV-2, we then test the potential to use local surveillance to recover underlying international infection dynamics.

## Methods

### Data

Travel testing in French Polynesia was conducted in two distinct phases, with some additional adjustments to protocols during each phase. The first phase, between 15^th^ July 2020 and 30^th^ April 2021, was the ‘COV-CHECK’ system, with travellers required to perform a PCR test less than 72 hours before departure as well as a self-test four days after arrival in French Polynesia [15]. Mandatory quarantine was added to this protocol on 20^th^ February 2021. A schematic of the process is shown in Figure 1A.

**Figure 1:**
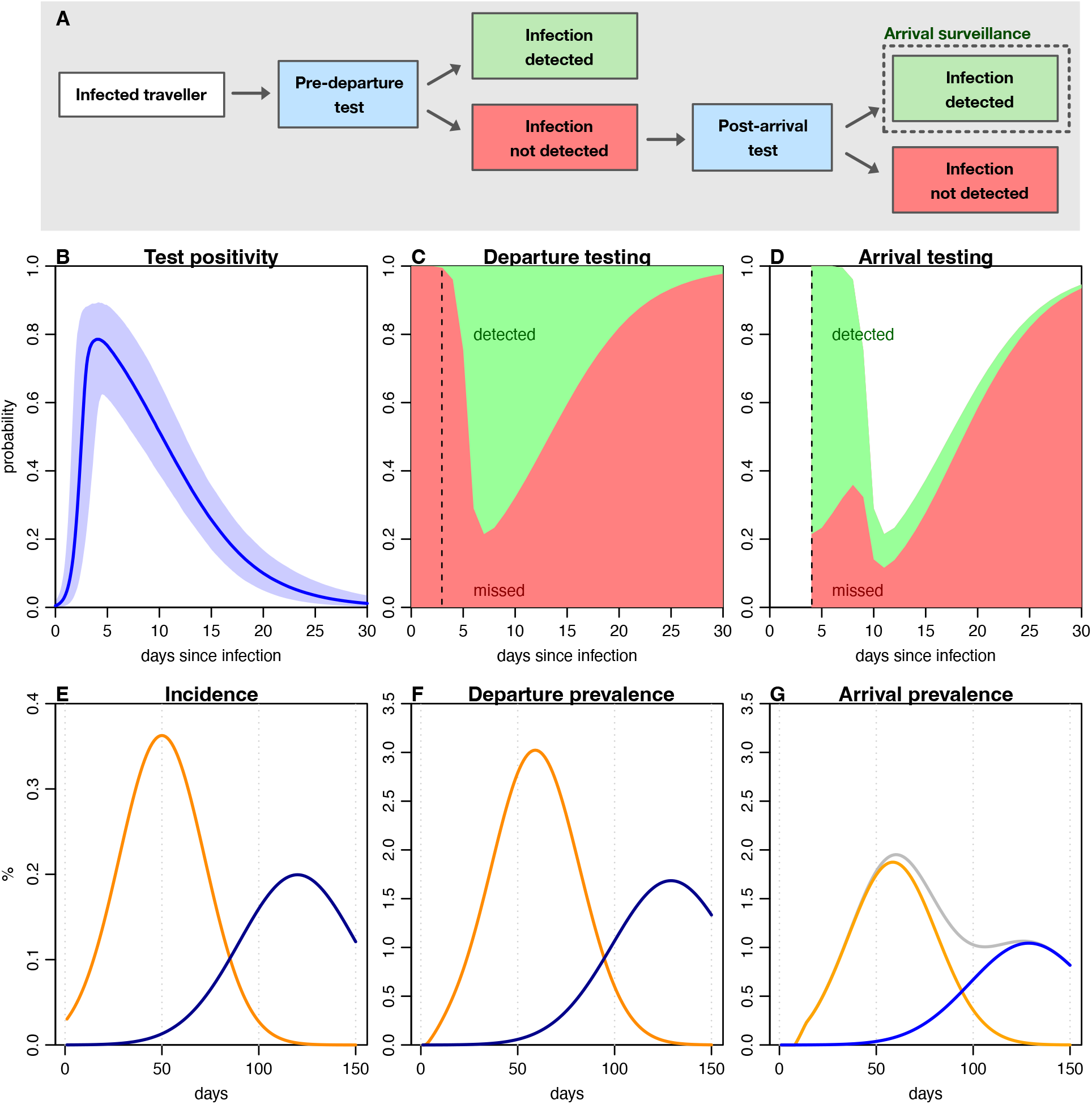
Impact of departure and arrival testing protocols on PCR prevalence. A) Possible outcomes for infected travellers in a scenario with pre-departure and post-arrival testing. B) Probability an individual will test PCR positive at different points since infection, based on self-tested asymptomatically tested participants [Hellewell et al]. Line shows median, shaded region 95% CrI. C) Probability infected traveller will be detected by a pre-departure test, in scenario where test conducted 2 days before departure (dashed line). D) Probability infected traveller will be detected by a post arrival test conducted 4 days after arrival (dashed line), assuming no local acquisition of infection. E) Illustrative epidemic showing proportion of population newly infected per day with two different variants. F) Larger measured prevalence in pre-departure testing corresponding to incidence curves in (E), based on positivity probability in (C). G) Measured prevalence in post-arrival testing, corresponding to incidence curves in (E), based on positivity probability in (D). Grey line shows cumulative prevalence.

The second phase started on 1^st^ May 2021 with the option of taking either an antigen test within 48 hours of departure or a PCR test within 72 hours. Moreover, testing was performed on the day of arrival in French Polynesia [16] by nurses until 12^th^ August 2021, then using the self-test COV-CHECK protocol until 27^th^ December 2021, and finally reverted to nurses until the end of the surveillance period.

Unvaccinated individuals (mostly children) had additional self-tests on day 4 and day 8 until 19^th^ January 2022, then on day 2 and 5. As these repeat tests did not systematically have accompanying dates in the dataset, and hence it was not possible to identify which test was conducted on day 0, we omitted these repeated test results (7809 of 112945 total test results that had a recorded country of origin) from the analysis. After 28^th^ December 2021, departing travellers could take either an antigen test within 24 hours of travel or a PCR test within 72 hours. Throughout the pandemic, subset of arrival PCR tests were sequenced to identify imported variants, with sequencing activity typically focused in the early stages of each wave to detect initial arriving infections.

### Travel testing model

If testing is implemented at both departure and arrival, then infections detected among arriving travellers only reflect a subset of all the infected individuals who attempted to travel (Figure 1A). Analysis of repeated PCR self-testing in a healthcare worker cohort [17], found that individuals can test positive for several weeks, with a peak in detection around 5 days post infection (Figure 1B). Departure testing is therefore most likely to prevent the most detectable infections from travelling (Figure 1C), which means if arrival testing is conducted shortly after arrival, the proportion of infections detected will be distributed differently to the departure tests (Figure 1D).

For a given incidence of new infections in a country of departure, measured prevalence among departing and arriving travellers could therefore be considerably different, with overall arrival prevalence masking underlying dynamics in countries of departure (Figure 1E–G).

To estimate the probability of detection at departure, we used the median estimate of testing PCR positive from a cohort study of self-tested healthcare workers [17], taking the first D=30 days post-infection in our analysis as the probability was negligible after this point. We assume PCR positivity post-infection for the wildtype variant is representative of positivity for subsequent variants [22]. As pre-departure PCR testing had to be within 72 hours of travel, in our baseline scenario we assumed travellers performed a PCR test 2 days before departure, with 1 day travel time to French Polynesia (i.e. 3 days between test and arrival). Sensitivity analysis to this assumption is shown in the Supplementary Materials (Figure S1).

The prevalence estimation model was defined as follows. Let *p*_*i*_ be the probability that an infected individual will test positive on day *i* after infection and *x*_*j*_ be the number of individuals infected on day *j*. Then the number of infected travellers who would be detected in a departure test on day *k* is given by:

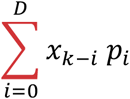

If pre-departure tests are conducted *t*_*pre*_ days before arrival, then the number of infected travellers who would go undetected and arrive on day *k + t*_*pre*_ is given by:

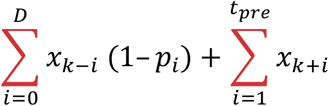

If post-arrival tests are conducted *t*_*post*_ days after arrival, then the number of infected arriving travellers who would test positive on day *k + t*_*pre*_ *+ t*_*post*_ post-infection is given by:

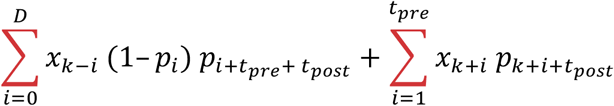

To calculate the scaling factor to convert arrival prevalence to an estimate of departure prevalence, we assumed stable epidemic incidence at departure (i.e. x_i_ = x_j_ for *i≠j*) and no infections post arrival (i.e. infection prevalence much lower at arrival destination versus departing country). We first define the probability that a departing infected individual would be *i* days post-infection and go undetected:

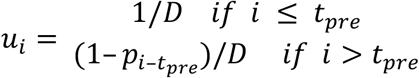

We then define the probability that an arriving infected individual would be *i* days post-infection and test positive at arrival, assuming that no infection occurred post-arrival while waiting to perform the test:

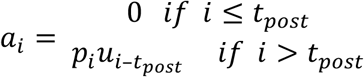

Next, we calculate the probability distribution for days post-infection among individuals who were detected at arrival, and normalise by probability of detection *i* days post-infection, then sum over this distribution to obtain the expected ratio of arriving infections per infection detected at arrival:

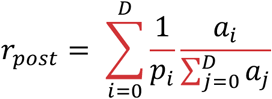

The ratio of infected individuals at point of arrival to those that would in theory have been detectable *t*_*post*_ days later can be defined as follows for each *i* day post-infection:

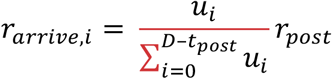

The ratio of departing infections to detected arriving infections is therefore given by

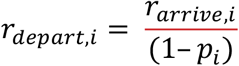

and hence the ratio of departure prevalence to arrival prevalence is calculated as:

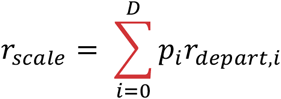

If *n*_*i*_ is the number of arrival tests performed in week *i* and *y*_*i*_ is the number of tests that were positive, then the proportion of individuals in this group who would have tested positive at departure was estimated as *r*_*scale*_*y*_*i*_ */ n*_*i*_. As shown in Figure 2, scaling measured arrival prevalence by *r*_*scale*_ could recover simulated prevalence across a range of scenarios, especially if there was a delay of several days between the pre-departure and post-arrival tests.

**Figure 2:**
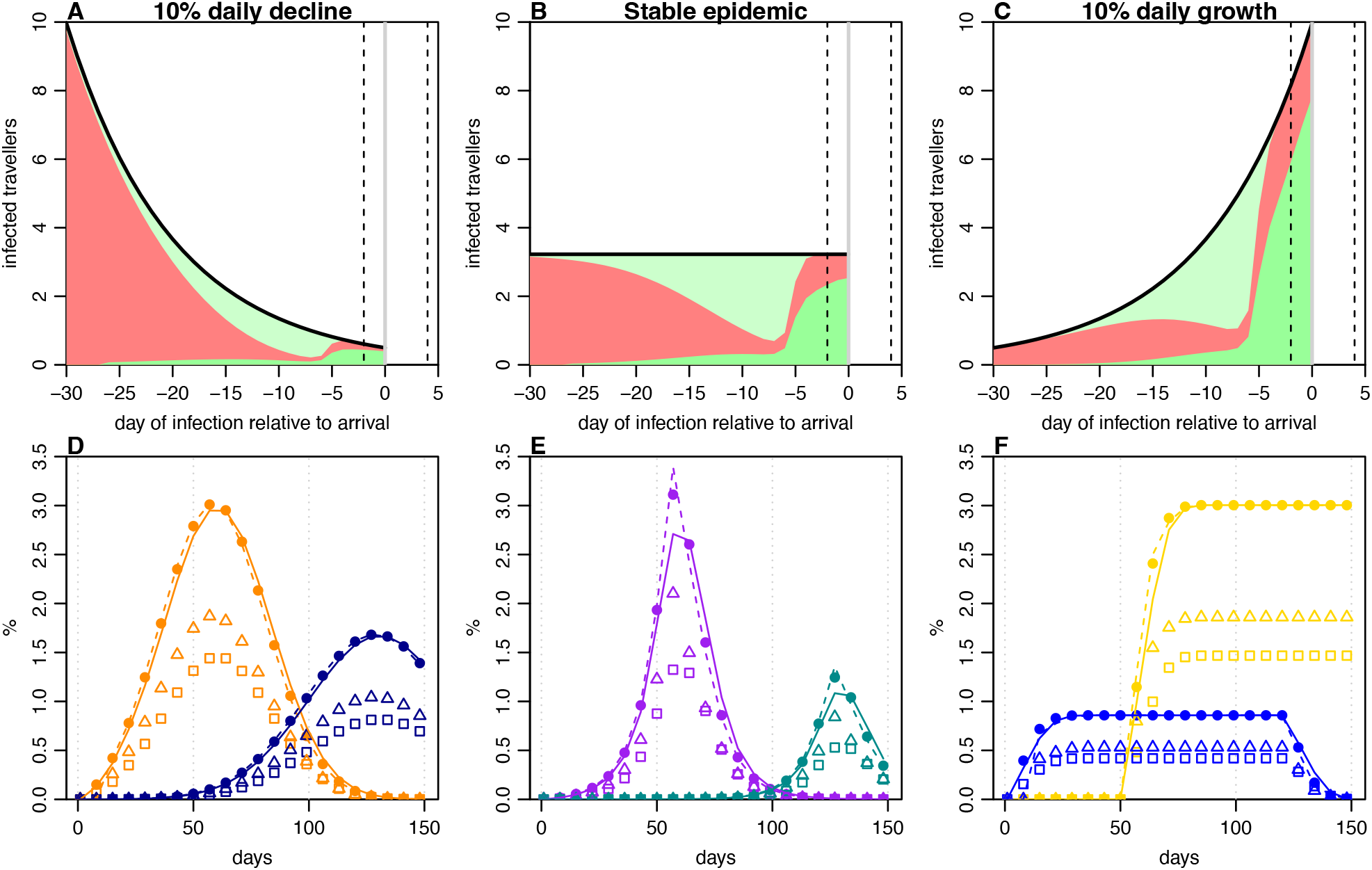
Observation and estimation of infection prevalence under different epidemic dynamics. A) Proportion of 100 infected travellers detected relative to infection time, in a scenario with incidence declining 10% per day, and testing 2 days before departure, with one day in transit, and 4 days after arrival (dashed lines). Light green, travellers detected pre-departure; dark green, travellers detected post-arrival; red, travellers missed. B) Scenario in a stable epidemic, i.e. 0% daily change in incidence. C) Scenario in epidemic with 10% daily growth. D) Reconstruction of simulated epidemics from arrival testing data. Solid points, ‘true’ prevalence at departure; squares, measured prevalence at arrival in scenario with test 1 day before departure and on day of arrival, with solid line showing estimated departure prevalence from these data; triangles, measured prevalence at arrival in scenario with test 3 days before departure and 4 days post-arrival, with dashed line showing estimated departure prevalence from these data. E–F) Reconstruction as described in (D) under different assumed epidemic dynamics.

### Prevalence model

To estimate daily departure prevalence, we used a generalised additive model (GAM) [18] using a binomial family and number of tests and estimated departure positives. We estimated pre and post 1^st^ May 2021 separately, as arrival testing shifted from day 4 to day 0 at this point, and hence the scaling factor also changed. To convert infection prevalence into an estimate of incidence, we divided weekly prevalence by the mean duration of positivity 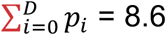 days, and assumed this corresponded to incident positivity 9 days earlier. This approximation is further explored in Figure S2.

We generated a bootstrap 95% prediction interval for cumulative incidence by simulating 1000 times from the fitted GAM using observed weekly intervals and testing numbers, calculating cumulative incidence, then estimating the median and central 95% values over time. To compare with seroprevalence data, we assumed a 17 day delay to seroconversion [19]. Model code and data are available at: https://github.com/institutlouismalarde/covid-travel-testing

### Ethical considerations

Communication of data from the surveillance system was approved by Comité d’ethique de la Polynésie française (ref Avis n°90 CEPF 15_06_2021). Secondary data analysis was approved by the LSHTM Observational Research Ethics Committee (ref 28129).

## Results

The effectiveness of travelling screening based on biological outcomes, such as symptoms or test positivity, will depend on how long ago travellers were infected. In turn, this will depend on the dynamics of the epidemic in countries of departure [20]. During a growing epidemic, individuals are more likely to have been infected recently, whereas infections will generally be older in a declining epidemic. In situations where test positivity is influenced by the phase of the epidemic, it is often necessary to reconstruct infection dynamics using computationally intensive latent processes [21]. However, we found that the presence of departure testing changes the distribution of infection timings among arrivals, as the individuals most likely to be detectable are least likely to travel (Figure 2A–C). As a result, most arriving infections will have been infected within a narrower window that those detected at departure. This reduced the impact of epidemic phase on arrival positivity and allowed for reconstruction of departure prevalence from arrival data with a single multiplier calculated from PCR positivity curves and the specific departure and arrival testing protocol. Our approach could therefore generate reliable estimates for simulated departure prevalence dynamics, particularly if there was a gap of several days between the departure and arrival test, which reduced the variance of the time-since-infection for arrivals (Figure 2D–F).

To show how this approach can be used mid-pandemic, we analysed traveller data collected as part of the COVID-19 response in French Polynesia [15]. Between July 2020 and March 2022, more than 222,000 traveller tests were conducted as part of an arrival screening programme (Figure 3A). During the first phase of the surveillance strategy, travellers performed a self-test (COV-CHECK protocol) four days after arrival as well as a PCR test within 72 hours of departure. Faced with novel variants, mandatory quarantine was added to the protocol in February 2021. Between May 2021 and March 2022, testing was performed on the day of arrival first by nurses until 12^th^ August 2021, then using the self-test COV-CHECK protocol until 27^th^ December 2021, and again by nurses until the end of the surveillance period. During 2021, the option was also added for an antigen test within 48 hours of departure rather than a PCR test within 72 hours.

**Figure 3:**
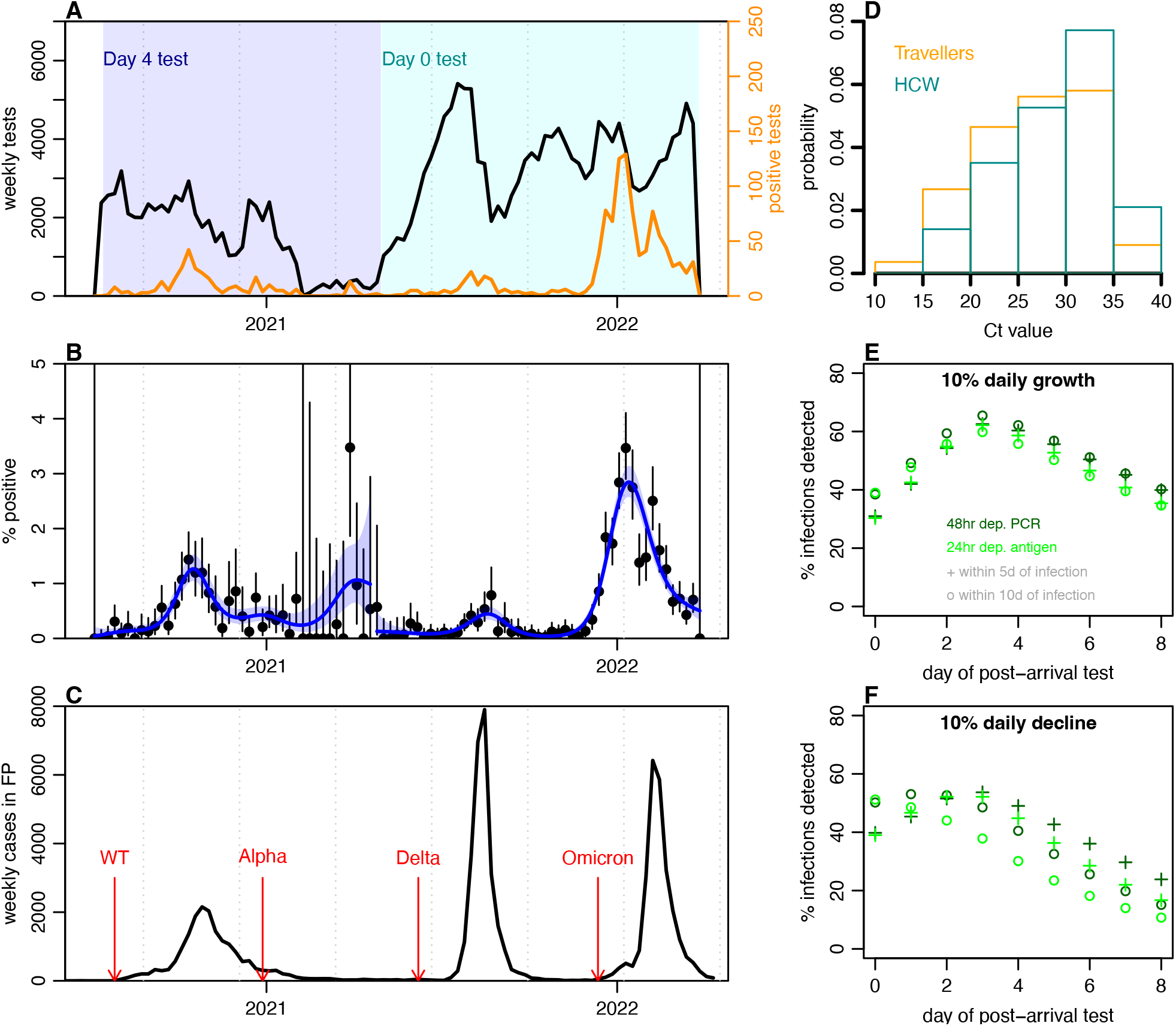
Arrival testing in French Polynesia, July 2020 to March 2022. A) Testing data and changes in main protocols over time. Initially travellers were tested 4 days post-arrival (d4), with additional quarantine introduced on 20^th^ February 2021; later vaccinated travellers were tested on day of arrival (d0), with additional testing on day 4 and 8 for non-vaccinated individuals. Black line, number of tests performed; orange line, number of positive tests. B) Percent of tests that were positive over time, with lines showing binomial CI and blue line showing generalised additive model (GAM) fit with shaded 95% CI. C) Local COVID-19 cases reported in French Polynesia, with arrows showing first detection of different variants among travellers. D) Distribution of Ct values among positive arrivals into French Polynesia (orange bars) and routinely tested UK healthcare workers (cyan bars). E) Estimated percent of infections that would be detected early on (i.e. within 5 or 10 days of infection) under different travel testing protocols in a growing epidemic. F) Estimated percent of infections detected in a declining epidemic.

There were 1341 positive arrival tests in the dataset analysed, with considerable variation in weekly prevalence over the course of the pandemic (Figure 3B). There were three main COVID-19 waves in French Polynesia – caused by wildtype, Delta and Omicron – with Alpha detected among travellers without leading to widespread local transmission (Figure 3C). Among arriving travellers, 15% of measured Ct values were below 20 (Figure 3D), with a distribution that was lower than has previously been observed in self-testing of healthcare workers [17]. This is to be expected given travellers will typically be earlier in their infection at the point of the arrival test, as noted above (Figure 2). Such a pattern is also consistent with previous reports of shedding being higher than usual among positive arriving travellers [22]. Based on protocols implemented, we estimated that arrival PCR testing at day 4 would have detected around 60% of infected individuals within the first 10 days of their infection if an epidemic in the departure location was growing at 10% per day (Figure 3E), and around 40% during an epidemic that was declining 10% per day (Figure 3F). This illustrates the value of delayed arrival testing during rising SARS-CoV-2 transmission.

The majority of tested arrivals into French Polynesia had either the USA or metropolitan France (with a few hours transit via the USA or Canada) as the origin for their trip (Figure 4A–B). This was consistent with reported residency data on immigration forms, with 90% of arrivals in 2021 from these two countries [23]. We found that measured prevalence at arrival among travellers from the USA or France anticipated observed case dynamics in the two countries (Figure 4C–D), showing the value of traveller testing as a leading indicator: peaks in prevalence occurred shortly before peaks in reported cases, as would be expected due to delays in symptom onset, symptomatic testing and reporting in the country of origin. Adjusting for traveller testing protocols as outlined in Figure 1, we estimated a peak infection prevalence at departure of 2.8% (2.3-3.6%) in France and 1.1% (0.81-3.1%) in the USA in late 2020/early 2021, with prevalence of 5.4% (4.8-6.1%) and 5.5% (4.6-6.6%) respectively estimated for the Omicron BA.1 waves in early 2022. For context, PCR test positivity in the ONS community infection survey in the UK peaked at around 2% during the Alpha wave in early 2021 and around 7% during the BA.1 wave in late 2021/early 2022 [7].

**Figure 4:**
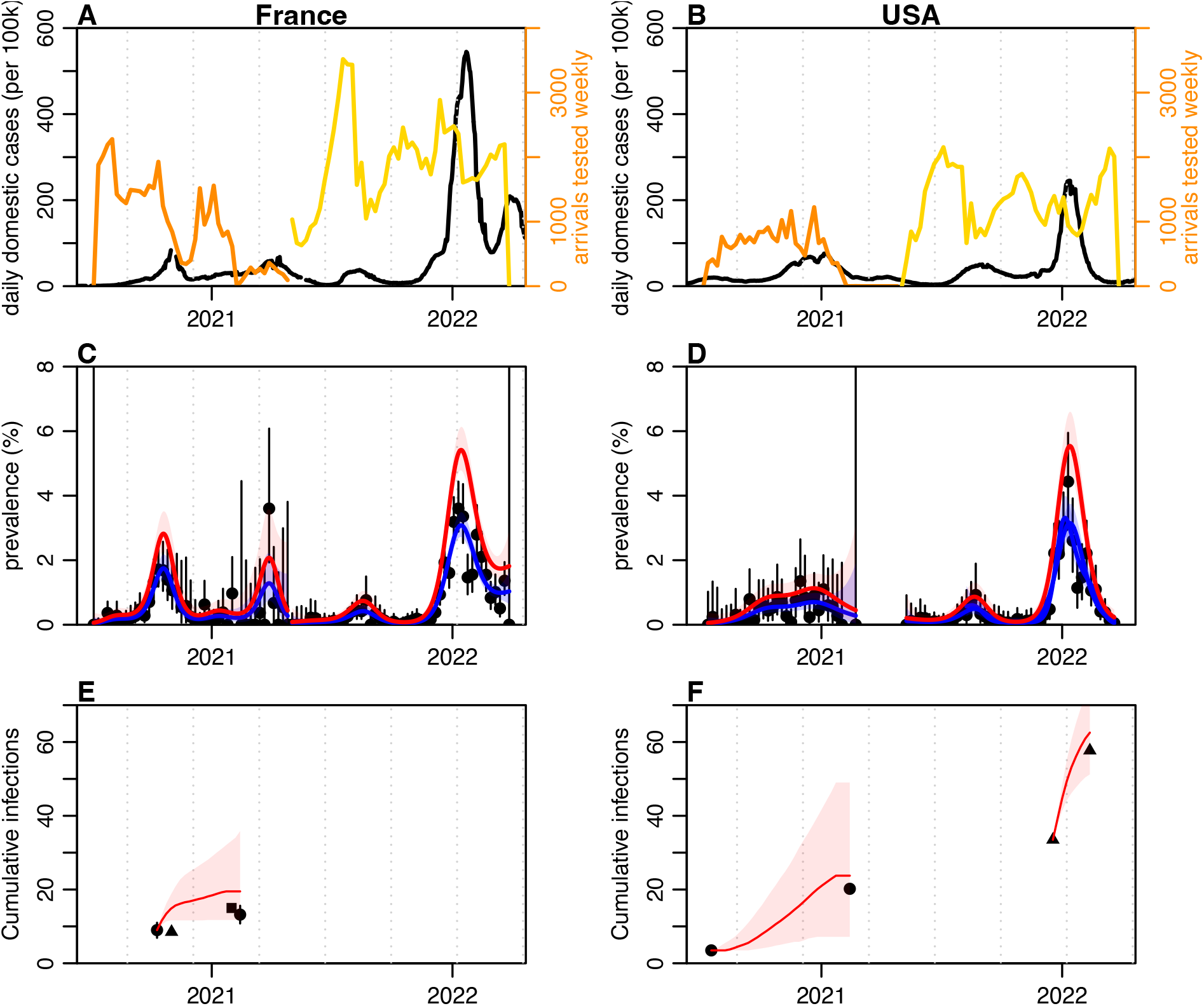
Reconstruction of infection dynamics in France and USA from arrival testing data in French Polynesia. A) Number of arrivals from France tested per week. Orange, tests performed at day 4 after arrival; yellow, tests performed at day of arrival. Black line shows domestic cases reported in France. B) Arrival testing and domestic case data for USA. C) Observed and estimated prevalence among arrivals from France, shown by black dots with 95% binomial CI; blue line and shaded region, GAM fit to these data and 95% CI; red line and shaded region, estimated prevalence at departure in France. D) Observed and estimated prevalence among arrivals from the USA. E) Comparison of estimated cumulative infections and observed seroprevalence in France. Black dots, observed national seroprevalence in France in October 2020 and February 2021 [26]; black triangle, observed national seroprevalence in November 2020 [27]; black square, estimated proportion infected by January 2021 [28] red line, cumulative incidence derived from red line in (E), shifted to match initial value of black line; shaded region, bootstrap 95% confidence interval. F) Estimated cumulative infections and observed seroprevalence in USA. Black dots, observed seroprevalence in July 2020 and May 2021 [29]; triangles, observed seroprevalence in December 2021 and February 2022 [30]; red lines, estimated cumulative incidence over same periods, shifted to match initial values.

To further assess the ability of arrival screening to estimate international SARS-CoV-2 dynamics, we converted prevalence at departure (i.e. current positivity) into an estimate of incidence (i.e. new daily infections), and compared cumulative incidence over the same period as repeated serological surveys in France and the USA. In France our estimate of cumulative incidence in late 2020 was slightly larger than implied in serological studies (Figure 4E). There are several potential explanations for this discordance. The volume of travel from France declined substantially during that wave, which may have affected representativeness of travellers, as risk-averse individuals could have been less likely to travel, and risk-taking individuals more likely to be infected in transit. As testing was conducted on day 4 during this period and there was a COVID-19 wave in French Polynesia in late 2020, there is also the possibility of infection post-arrival. In contrast, we found that our estimates for the USA closely matched increases in seroprevalence during epidemic waves in both late 2020 and late 2021 (Figure 4F).

## Discussion

Our analysis shows that it is possible to obtain detailed international epidemiological insights from arrival testing protocols that were designed predominantly as control measure to reduce risk of onwards local transmission. In French Polynesia, further data linkage was limited to protect traveller privacy. However, insights could be expanded by routinely reporting data on epidemiologically relevant values such as age and whether travelling in a group, which would allow adjustment for potential biases when estimating prevalence in countries of departure.

Observed infection prevalence at arrival in French Polynesia was consistent with broad ranges observed in other, smaller-scale studies. Among arrivals into Alaska from June to November 2020, 0.8% were positive [12]; Infection prevalence at arrival was 1.0% in Toronto, Canada during September and October 2020 [14] and 1.5% in Alberta, Canada in November 2020 [13]. However, travellers in these studies did not have their country of origin reported, or information on departure testing protocol, limiting the ability of these datasets to provide comparable information on likely prevalence in countries of departure.

There are some additional limitations to our proof-of-concept analysis. The testing protocols in French Polynesia enabled estimation of departure prevalence using a simple scaling adjustment, with little bias from epidemic phase (Figure 2E–G). In the absence of departure testing, such methods would no longer be reliable, and arrival testing would need to be analysed in the context of changing epidemic dynamics, requiring more complex inference methods such as Gaussian process models [21]. We also assume that PCR positivity over time for the wildtype variant is representative of subsequent variants, based on similarities observed in cohort studies [24]. If positivity were to vary substantially, then this would need to be account for with different adjustments based on dominant variants in countries of departure.

During the initial phase of COVID-19 pandemic, testing among departing and arriving travellers provided valuable indications of the true extent of infection [3], which were in turn used to estimate crucial metrics such as infection fatality risk [25]. Understanding levels of community infection has been challenging both when infections are rising, with the epidemic outstripping symptomatic surveillance capacity, as well as in the later stages of the COVID-19 pandemic, with countries rolling back test availability for symptomatic cases. In the UK, studies such as REACT-1 and ONS have provided continuity in local understanding of community infections, but such data streams have been extremely rare globally. However, our analysis shows that routine traveller testing can enable similar insights at an international scale, using protocols that were already being implemented. It therefore provides a proof-of-concept for ongoing COVID-19 management and future pandemic planning, showing that systematic collection of testing data with minimal linkage can enable real-time estimation of underlying epidemic dynamics in multiple countries. Moreover, deployment of such methods in multiple locations – such as key global travel hubs – would allow for synthesis of prevalence estimation across datasets, narrowing uncertainty and greatly expanding the network of countries covered.

## Data Availability

Model code and data are available at: https://github.com/institutlouismalarde/covid-travel-testing

## References

1. Koelle, K., Martin, M. A., Antia, R., Lopman, B. & Dean, N. E. The changing epidemiology of SARS-CoV-2. Science 375, 1116–1121 (2022).

2. Tuite, A. R. et al. Estimation of Coronavirus Disease 2019 (COVID-19) Burden and Potential for International Dissemination of Infection From Iran. Ann. Intern. Med. 172, 699–701 (2020).

3. Imai, N. et al. Report 2: Estimating the potential total number of novel Coronavirus cases in Wuhan City, China. 6 (2020).

4. Jit, M. et al. Estimating number of cases and spread of coronavirus disease (COVID-19) using critical care admissions, United Kingdom, February to March 2020. Eurosurveillance 25, (2020).

5. Kalish, H. et al. Undiagnosed SARS-CoV-2 seropositivity during the first 6 months of the COVID-19 pandemic in the United States. Sci. Transl. Med. 13, eabh3826 (2021).

6. Chapman, L. A. C. et al. Unexposed populations and potential COVID-19 hospitalisations and deaths in European countries as per data up to 21 November 2021. Eurosurveillance 27, 2101038 (2022).

7. Office for National Statistics. Coronavirus (COVID-19) Infection Survey (2022). Available from: https://www.ons.gov.uk/peoplepopulationandcommunity/healthandsocialcare/conditionsanddiseases/bulletins/coronaviruscovid19infectionsurveypilot/previousReleases

8. Riley, S. et al. Resurgence of SARS-CoV-2: Detection by community viral surveillance. Science 372, 990–995 (2021).

9. Hall, V. J. et al. SARS-CoV-2 infection rates of antibody-positive compared with antibody-negative health-care workers in England: a large, multicentre, prospective cohort study (SIREN). The Lancet 397, 1459–1469 (2021).

10. Ritchie, H. et al. Coronavirus Pandemic (COVID-19). Our World Data (2020).

11. Grout, L. et al. Failures of quarantine systems for preventing COVID-19 outbreaks in Australia and New Zealand. Med. J. Aust. 215, 320–324 (2021).

12. Ohlsen, E. C. et al. Airport Traveler Testing Program for SARS-CoV-2 — Alaska, June–November 2020. MMWR Morb. Mortal. Wkly. Rep. 70, 583–588 (2021).

13. Lunney, M. et al. COVID-19 infection among international travellers: a prospective analysis. BMJ Open 11, e050667 (2021).

14. Goel, V. et al. COVID-19 international border surveillance at Toronto’s Pearson Airport: a cohort study. BMJ Open 11, e050714 (2021).

15. Aubry, M. et al. Self-collection and pooling of samples as resources-saving strategies for RT-PCR-based SARS-CoV-2 surveillance, the example of travelers in French Polynesia. PLOS ONE 16, e0256877 (2021).

16. Aubry, M & Cao-Lormeau, V.-M.. Perspective on the Use of Innovative Surveillance Strategies Implemented for COVID-19 to Prevent Mosquito-Borne Disease Emergence in French Polynesia. Viruses (2022).

17. Hellewell, J. et al. Estimating the effectiveness of routine asymptomatic PCR testing at different frequencies for the detection of SARS-CoV-2 infections. BMC Med. 19, 106 (2021).

18. Wood SN. Generalized Additive Models: An Introduction with R. CRC Press (2006)

19. Borremans, B. et al. Quantifying antibody kinetics and RNA detection during early-phase SARS-CoV-2 infection by time since symptom onset. eLife 9, e60122.(2020)

20. Gostic, K. M., Kucharski, A. J. & Lloyd-Smith, J. O. Effectiveness of traveller screening for emerging pathogens is shaped by epidemiology and natural history of infection. eLife 4, e05564 (2015).

21. Abbott, S. & Funk, S. Estimating epidemiological quantities from repeated cross-sectional prevalence measurements. MedRxiv (2022)

22. Molero-Salinas, A. et al. High SARS-CoV-2 viral load in travellers arriving in Spain with a negative COVID-19 test prior to departure. J. Travel Med. 29, taab180 (2022).

23. Institut de la statistique de la Polynésie française. https://data.ispf.pf/themes/SystemeProductif/Tourisme/Details.aspx

24. Bouton, T. C. et al. Viral dynamics of Omicron and Delta SARS-CoV-2 variants with implications for timing of release from isolation: a longitudinal cohort study. MedRxiv (2022).

25. Verity, R. et al. Estimates of the severity of coronavirus disease 2019: a model-based analysis. Lancet Infect. Dis. 20, 669–677 (2020).

26. Santé Publique France. COVID-19: études pour suivre la part de la population infectée par le SARS-CoV-2 en France (2021). Available from: https://www.santepubliquefrance.fr/etudes-et-enquetes/covid-19-etudes-pour-suivre-la-part-de-la-population-infectee-par-le-sars-cov-2-en-france

27. DREES. 4 % de la population a développé des anticorps contre le SARS-CoV-2 entre mai et novembre 2020. Études et Résultats (2021).

28. Gallian, P. et al. SARS-CoV-2 IgG seroprevalence surveys in blood donors before the vaccination campaign, France 2020-2021. MedRxiv (2022)

29. Jones, J. M. et al. Estimated US Infection- and Vaccine-Induced SARS-CoV-2 Seroprevalence Based on Blood Donations, July 2020-May 2021. JAMA 326, 1400 (2021).

30. Clarke, K. E. N. et al. Seroprevalence of Infection-Induced SARS-CoV-2 Antibodies — United States, September 2021–February 2022. MMWR Morb. Mortal. Wkly. Rep. 71, 606–608 (2022).

