## Supplementary Materials for "Real-time surveillance of international SARS-CoV-2 prevalence using systematic traveller arrival screening"

### *Alternative departure protocol*

In our baseline scenario we assumed all travellers performed PCR tests within 2 days of departure. If we instead assume that all travellers performed antigen tests at departure within 1 day of travel, which they were eligible to do after May 2021, we estimated 4.2% (3.7-4.7%) and 4.5% (3.6-5.1%) during the BA.1 waves in France and the USA respectively (Figure S1).

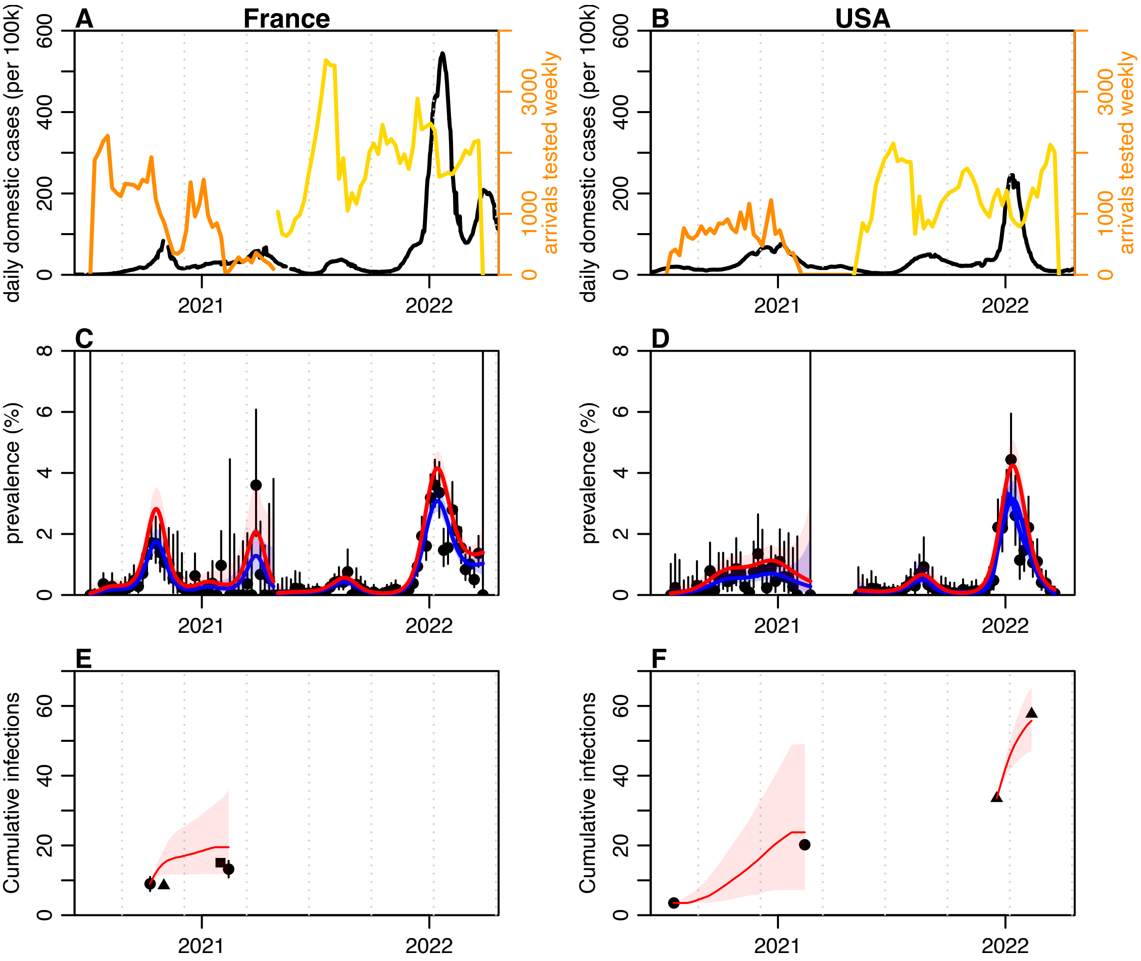


*Figure S1: Reconstruction of infection dynamics in France and USA from arrival testing data in French Polynesia, assuming antigen tests at departure. A) Number of arrivals from France tested per week. Orange, tests performed at day 4 after arrival; yellow, tests performed at day of arrival. Black line shows domestic cases reported in France. B) Arrival testing and domestic case data for USA. C) Observed and estimated prevalence among arrivals from France, shown by black dots with 95% binomial CI; blue line and shaded region, GAM fit to these data and 95% CI; red line and shaded region, estimated prevalence at departure in France. D) Observed and estimated prevalence among arrivals from the USA. E) Comparison of estimated cumulative infections and observed seroprevalence in France. Black dots, observed national seroprevalence in France in October 2020 and February 2021; black triangle, observed national seroprevalence in November 2020; black square, estimated proportion infected by January 2021; red line, cumulative incidence derived from red line in (E), shifted to match initial value of black line; shaded region, bootstrap 95% confidence interval. F) Estimated cumulative infections and observed seroprevalence in USA. Black dots, observed seroprevalence in July 2020 and May 2021; triangles, observed*

*seroprevalence in December 2021 and February 2022; red lines, estimated cumulative incidence over same periods, shifted to match initial values.*

***Reconstruction of cumulative incidence***

In our baseline scenario, we reconstruct cumulative incidence in Figure 4 by scaling prevalence by the mean duration of positivity. To explore the performance of this approximation, we simulated daily incidence (Figure S2A), converted this incidence to prevalence by convolving with probability of PCR positivity post-infection (Figure S2B). We then re-estimated incidence from these prevalence data using our baseline scaling approximation (red line in Figure S2C) as well as deconvolution based on the Moore-Penrose generalized inverse (blue line in Figure S2C). Estimated cumulative incidence is shown in Figure S2D, showing that the simple scaling approximation gives a reasonable reconstruction of cumulative incidence for plausible prevalence dynamics.

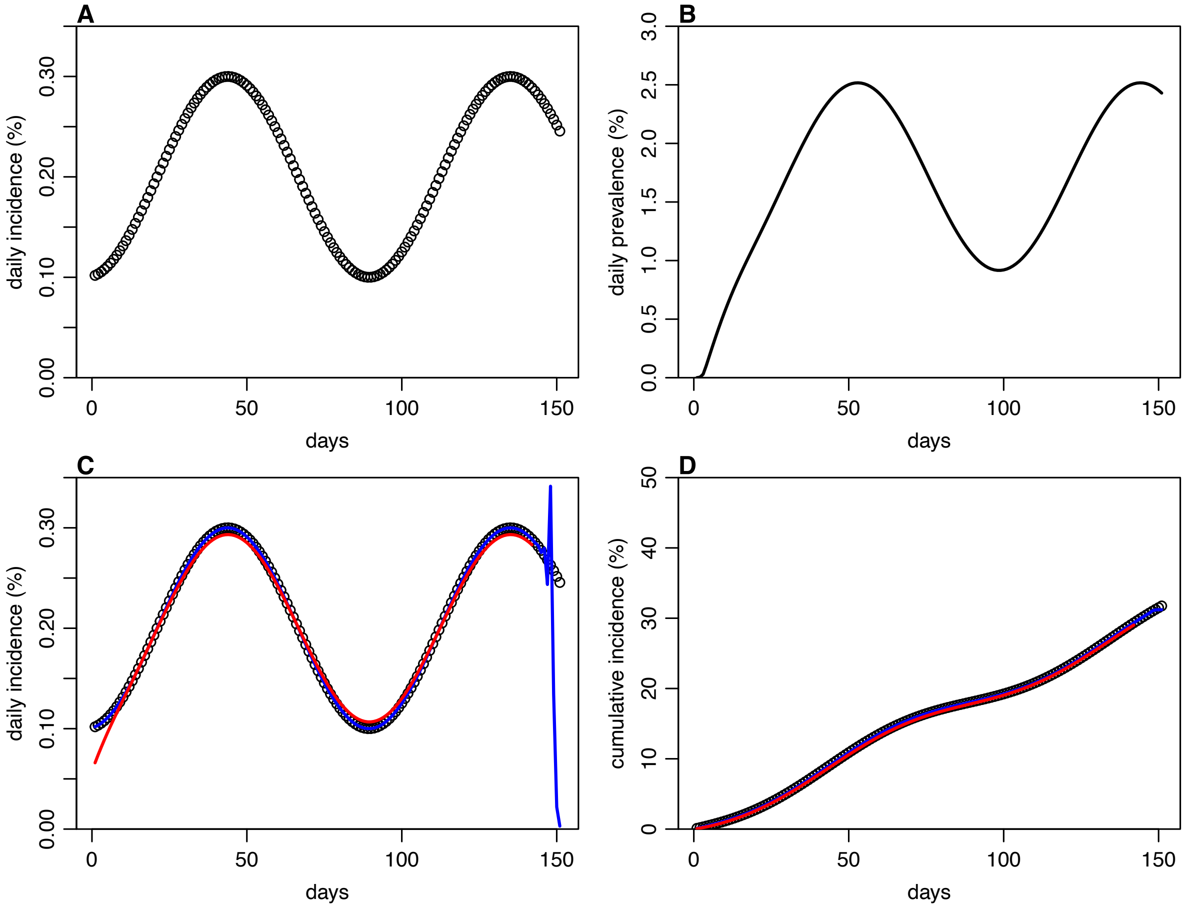


*Figure S2: Reconstruction of cumulative incidence under different estimated methods. A) Simulated daily incidence. B) Simulated daily prevalence based on the convolution of incidence in (A) and median probability of PCR positivity in Figure 1B. C) Reconstruction of incidence from prevalence in (B) using our baseline scaling approximation (red line) as well as deconvolution using the Moore-Penrose generalized inverse (blue line, showing numerical instability at end of time series). D) Comparison of simulated cumulative incidence (black dots), with estimated cumulative incidence using our baseline scaling approximation (red line) as well as deconvolution using the Moore-Penrose generalized inverse (blue line).*
